# Changes in the position of the hyoid bone in skeletal Class II children post-functional Activator therapy

**DOI:** 10.1101/2020.10.08.20208967

**Authors:** Cynthia Concepcion Medina, Hiroshi Ueda, Ryo Kunimatsu, Kotaro Tanimoto

## Abstract

**Introduction:** The hyoid bone is deeply involved in three important body functions: deglutition, phonation and respiration. Several studies have shown that changes in the position of the hyoid bone may influence in pharyngeal size with mandibular advancement, thus a forward positioned hyoid bone may be an indicator of wide upper airways.

**Objective:** To determine the changes in the position of the hyoid bone after functional appliance treatment.

**Subjects and methods:** 20 children (aged 9-13) that currently visit Hiroshima University Hospital actively undergoing FKO activator therapy volunteered for this study. Several lateral cephalometric radiographs were indicated and traced to assess hyoid bone position changes that might have occurred when actively complying with the FKO therapy, said radiographs were procured before active functional treatment (T0), during it (T1), and a year after continuous use of this appliance (T2). ANOVA tests were done to find statistical significance.

**Results:** The results of these tests were analyzed and compared; it was found that, the hyoid bone is at a lower position from the mandibular plane and Frankfurt horizontal plane after FKO treatment, also the mandible is more forward after activator therapy, bringing the hyoid bone forward with it thus widening the lowest section of the upper airways.

**Conclusion:** The FKO not only induces the proper development of the mandible, it also potentially advances the position of the hyoid bone, thus affecting positively in the opening of airways providing an improvement in the children’s breathing functions.

- Authors report no conflict of interest.

**Funding:** This research did not receive any specific grant from funding agencies in the public, commercial, or not-for-profit sectors.

**Conflicts of interest/Competing interests:** the authors have no conflicts of interest to declare.

**Ethics approval:** This research has been approved by the Ethics Review Committee of Hiroshima University (No. E – 56).

**Consent to participate:** All subjects needed to have provided informed consent from the parent or guardian prior all evaluations. This study has followed the guidelines stated in the Helsinki Declaration for clinical investigations.

**Consent for publication:** All subjects have provided informed consent from the parent or guardian to allow publication of acquired data.

## Introduction

The position of the hyoid bone has received considerable attention when it concerns to the relation of its placement and the collapsibility of the upper airways. Studies have shown that in various samples of several population groups, that the changes in the hyoid bone position seem to be related to the changes in the mandibular position and other facial structures in general (Amayeri *et al*.,2014).

Bibby and Preston, (1981), Trenouth, (1999) and Ferraz *et al*., (2006) previous studies have mentioned the importance of the fact that the high relevance of the hyoid bone lied in its unique anatomic relationship, when examined in response to mandibular advancement in subjects with mild and moderate Obstructive Sleep Apnea (OSA). A previous study (Battagel *et al*., 1999) showed that in the protruded mandible the hyoid bone became closer to the mandibular plane in the same time it got a more upward position. For this study, we examined skeletal Class II children and compared the results to those of skeletal Class I children to determine the differences in position of this structure in different craniofacial patterns and in different time points, especially after functional activator treatment in skeletal Class II children.

## Subjects and methods

Bearing in mind the anatomical differences that divide facio-skeletal patterns into Class II (SNA,SNB, and ANB angles), the participants of this study were evaluated according to the characteristics that the literature consensus agrees are skeletal Class I and Class II respectively (table I).

**Table I.** Summary of the skeletal relationship of the subjects for this study.

| | $\angle SNA^\circ$ (antero–<br>posterior position of the<br>maxilla) | $\angle SNB^\circ$ (antero–posterior<br>position of the mandible) | $\angle ANB^\circ$ (relationship between<br>the maxilla and the mandible) |
| --- | --- | --- | --- |
| **Control**<br>**group** | $81.3 \pm 0.3$ | $76.4 \pm 2.3$ | $4.8 \pm 1.2$ |
| **Activator**<br>**group** | $80.7 \pm 2.5$ | $74.6 \pm 0.9$ | $6.8 \pm 1.7$ |

Other inclusion/exclusion criteria for both groups included the absence of systemic diseases including bt not limited to

For the skeletal Class II children (Activator group), 20 children, 10 girls and 10 boys, aged 10.7 +/- 1.8 years old, for the skeletal Class I children (control group), 19 children, 13 boys and 6 girls, aged 10.8 ± 1.0 years, who attended the orthodontic clinic at Hiroshima University Hospital between 2014-2017 (table I), volunteered to participate in this study. All children needed to clear an inclusion criteria that indicated overall health with no previous history of sleep-related child breathing disorders. For the activator group, the subjects needed to be in the active stages of myofunctional activator treatment, for the control group, the orthodontic appliance used during active treatment was irrelevant as long as it did not have a direct influence on the patients’ skeletal growth nor the upper airways.

This study received the aproval by the Ethics Review Committee of Hiroshima University (No. E-56). All subjects needed to provide informed consent from their parents or guardian prior evaluation.

Using a lateral cephalometric radiograph to assess different anatomical points in relation to the position of the hyoid bone, noting the discrepancy between the two previously mentioned skeletal patterns in Japanese children. Furthermore, the data was collected in three points in time for the study group, T0,initial assessment; T1, with the activator inserted, and T2, final assessment after activator therapy, with a mean average of time of 8.3 ± 2.3 months elapsed in between stages. For the control group T0 and T2 were assessed, with a mean average time of 16 ± 2.6 months elapsed between stages.

The analysis that were included in this study incorporated the linear and angular measurements used in previous studies and depicted in Figure 1 (Bibby and Preston, 1981; Deljo *et al*., 2012; Jose *et al*.,2014).

**Figure 1.**
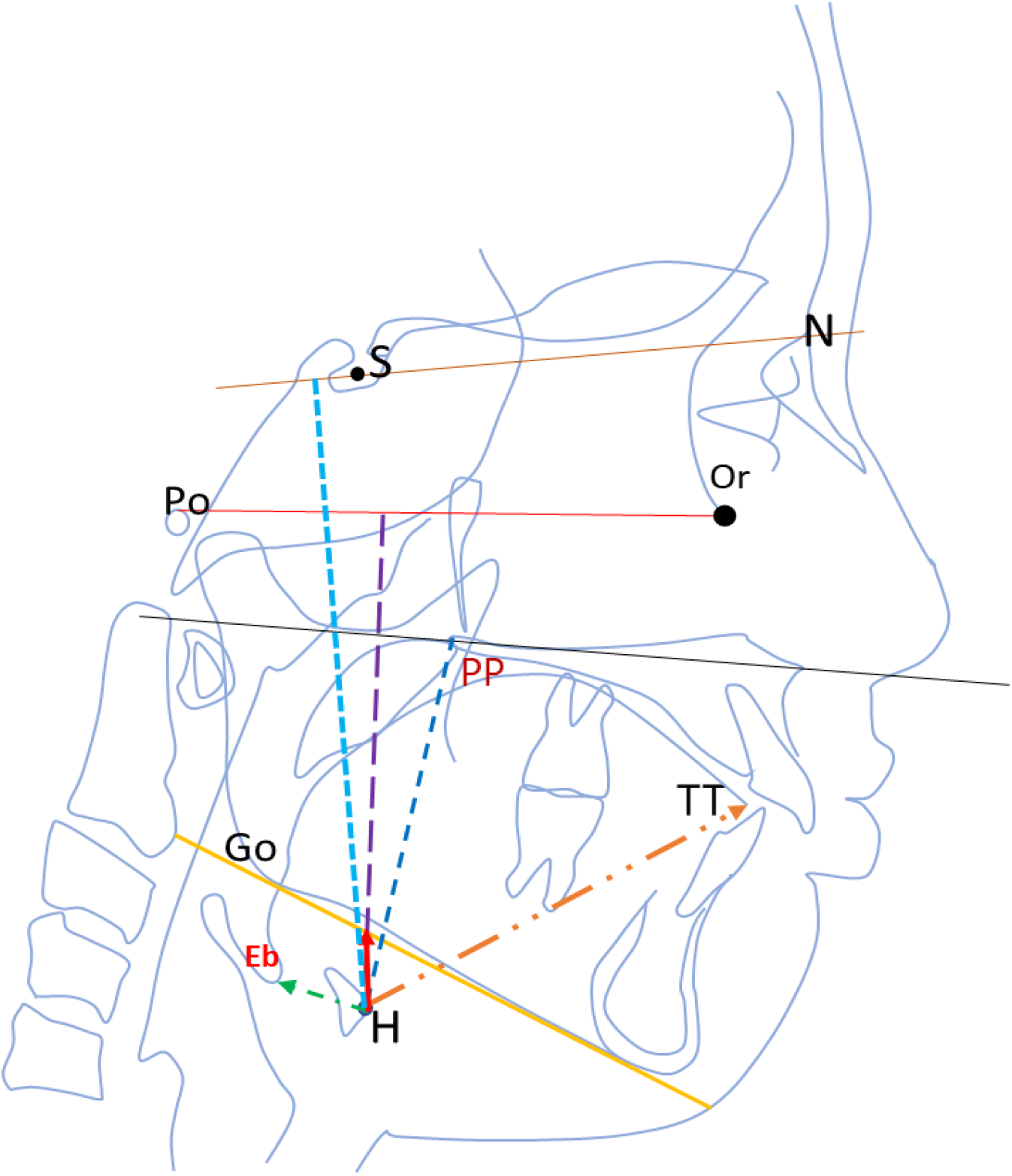
Linear and angular measurements for the position of the hyoid bone. SN-H perpendicular: linear distance along a perpendicular from H to the S-N plane. FH-H perpendicular: linear distance along a perpendicular from H to FH plane. H-MP: linear distance between H and MP. H-PP: linear distance between H and palatal plane. H-TT: linear distance between H and the tip of the tongue. H-Eb: linear distance between H and Eb (epiglottis) SH: the angle from nasion to sella to hyoidale. MPH the angle from gonion to gnathion to hyoidale. H-Eb-TT: the angle from base of epiglottis to tip of the tongue to hyoidale.

Previous studies have also remarked on the fact that there is a great variability on its position depending on even the slightest movement of the head. Therefore, they recommended to determine its position by using the cervical vertebra and the mandible as reference points as well the following measurement points:

### The hyoid triangle

The hyoid triangle allows determination of hyoid bone position in three directions and, since it is not dependent on a cranial reference plane, incorrectness that may stem from changes in head posture is minimized (Figure 2) (Bibby and Preston 1981).

**Figure 2:**
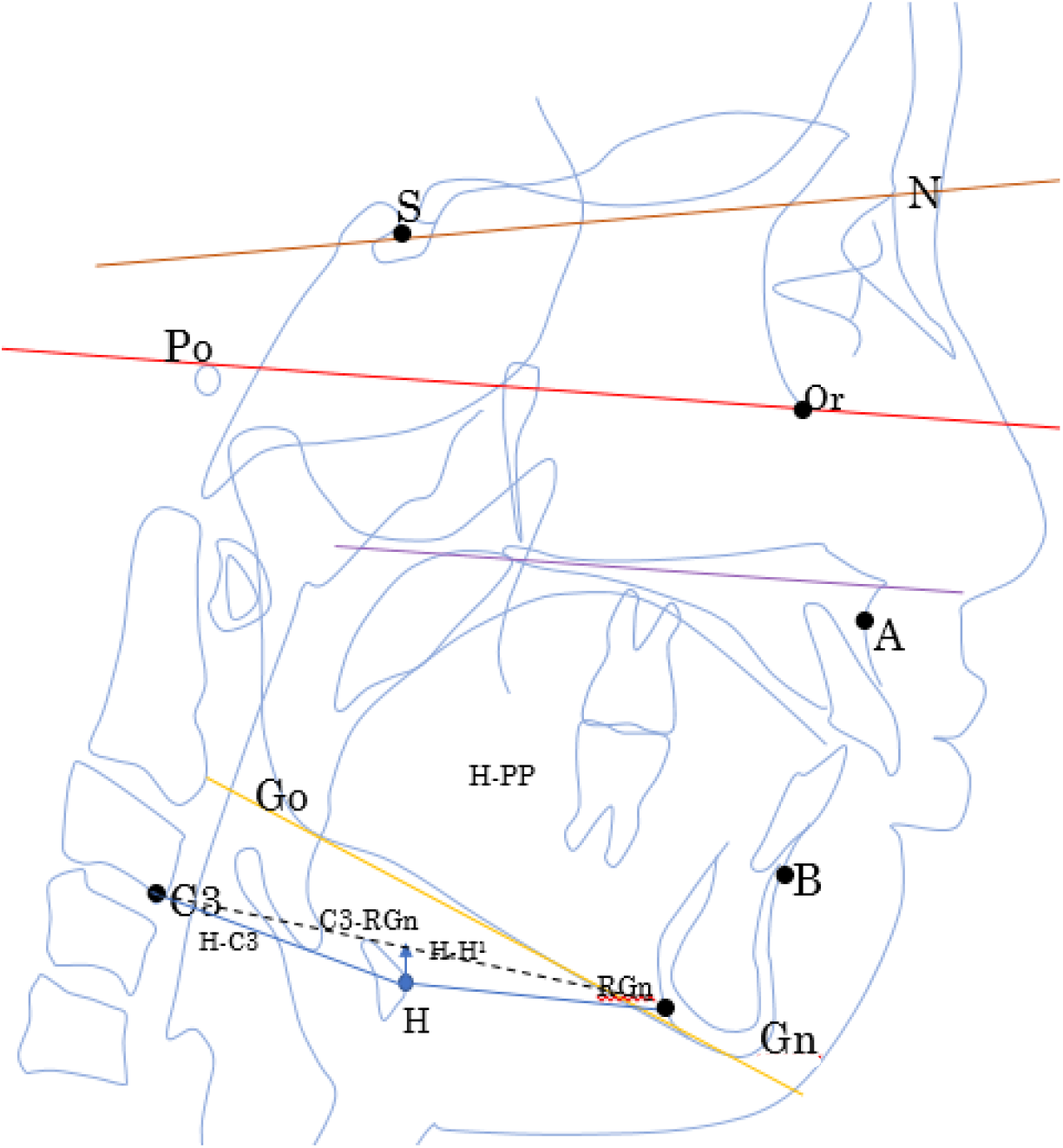
the hyoid triangle; C3-Rgn: linear distance between C3 and RGn C3-H: linear distance between H and C3. H-RGn: linear distance between H and RGn. H-H’: linear distance between H and a perpendicular to the C3- RGn line.

### Lower airway space

Based on the hypothesis that the more anteriorly positioned the hyoid bone is, the wider the lower section of the upper airways become, thus, the antero– posterior measurements in **Figure 3** were proposed and analyzed.

**Figure 3.**
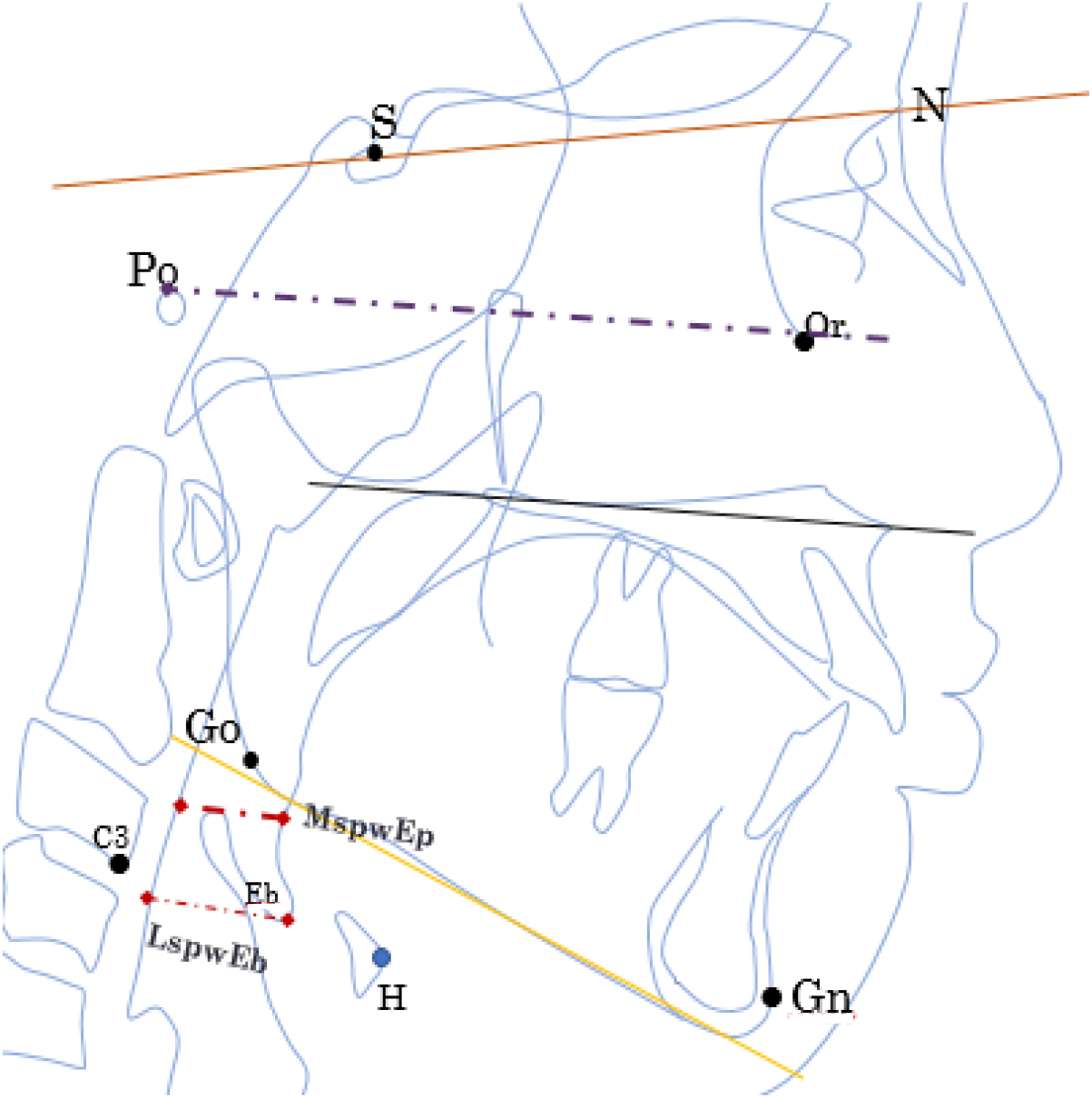
The lower airway space related to the hyoid bone. MspwEp: middle airway space assessed from the posterior border of the tongue body to the pharyngeal wall, touching the most superior point of the epiglottis in a line parallel to FH plane. LspwEb: lower airway space, evaluated from the base of the epiglottis to the pharyngeal wall in a line parallel to FH plane

All lateral cephalometric radiographs were taken in the natural head position by an experienced technician and manually traced by the first author, reliability was ensured by repeating random traces two weeks after the first trace to check for inconsistencies.

All radiographs were digitized using the free software ImageJ, U.S. National Institutes of Health, Bethesda, Maryland, USA, https://imagej.nih.gov/ij/, 1997-2016.

All statistical analyses were performed using Medcalc Statistical Software version 17.8.6 (Medcalc Software bvba, Ostend, Belgium; http://www.medcalc.org; 2017) and/or Microsoft Excel software, unless specified. Data are presented as mean ± standard deviation (sd); ANOVA tests were used to compare the difference between cephalometric results in both groups. A p-value of <0.05 was considered to indicate statistical significance.

## Results

After tracing, digitizing and assessing the chosen landmarks and measurement points, a statistical difference of P<0.01 was present in H–PP, H–FH and H– MP perpendicular measurements. In the sagittal plane, there is an increased distance of the hyoid bone from the anterior cranial base (H–SN perpendicular). Also, in relation to the mandible, it is shown that the hyoid bone takes a more inferior and anterior position when the activator is inserted, and this change is kept post–treatment. (table II)

**Table II.** Results for the linear assessment of the hyoid position.

| **Linear measurement**<br>**(cm)** | **Control**<br>**group** | **Activator group**<br>**(T0)** | **Activator group**<br>**(T1)** | **Activator group**<br>**(T2)** |
| --- | --- | --- | --- | --- |
| H–SN perpendicular | $8.99 \pm 0.5$ | $8.86 \pm 0.9^{**}$ | $9.16 \pm 0.7$ | $9.59 \pm 1.0^{**}$ |
| H–FH perpendicular | $7.03 \pm 0.6$ | $6.78 \pm 1.0^{**}$ | $7.34 \pm 0.6$ | $7.67 \pm 0.9^{**}$ |
| H–MP perpendicular | $0.81 \pm 0.4$ | $0.78 \pm 0.6$ | $0.48 \pm 0.3^{**}$ | $1.00 \pm 0.7^{**}$ |
| H–PP | $4.91 \pm 0.5$ | $4.94 \pm 0.7^{**}$ | $5.10 \pm 0.6$ | $5.36 \pm 0.6^{**}$ |
| H–TT | $4.77 \pm 0.4$ | $4.98 \pm 0.5$ | $4.98 \pm 0.3$ | $5.17 \pm 0.7$ |
| H–Eb | $1.45 \pm 0.3$ | $1.46 \pm 1.1$ | $1.23 \pm 0.2$ | $1.35 \pm 0.3$ |

**\*\*P<0.01**

When assessing the hyoid triangle, a larger distance was noticed between the mandible (from Rgn) and the third cervical vertebra (C3). Table II

The lower airway space especially measured from the base of the epiglottis to the pharyngeal wall (LspwEb) shows a significantly (P<0.01) increased sized when compared to initial data. (table III)

**Table III.** Comparison of the hyoid triangle among all groups.

| Cephalometric Variables | Control group | Activator group (T0) | Activator group (T1) | Activator group (T2) |
| --- | --- | --- | --- | --- |
| **C3-RGN (cm)** | 5.17 ± 0.7 | 5.38 ± 0.5 | 5.61 ± 0.6 | 5.70 ± 0.7 |
| **C3-H (cm)** | 2.95 ± 0.2 | 2.98 ± 0.4 | 3.10 ± 0.2 | 3.19 ± 0.3 |
| **H-RGN (cm)** | 2.66 ± 0.9 | 2.61 ± 0.4 | 2.63 ± 0.5 | 2.80 ± 0.5 |
| **H-H' (cm)** | 0.54 ± 0.3 | 0.56 ± 0.5 | 0.46 ± 0.3 | 0.68 ± 0.6 |

\*P< 0.05 \*\*P<0.01

**Table IV.** The lower section of the airways is wider when the hyoid position is influenced by the changes brought upon by the Activator.

| Cephalometric Variables | Control group | Activator group (T0) | Activator group (T1) | Activator group (T2) |
| --- | --- | --- | --- | --- |
| **MspwEp (cm)** | 1.12 ± 0.23 | 0.93 ± 0.34 | 1.01 ± 0.27 | 1.00 ± 0.26 |
| **LspwEb (cm)** | 1.52 ± 0.20 | 1.32 ± 0.22 | 1.45 ± 0.24 | 1.45 ± 0.29 |

\*P< 0.05 \*\*P<0.01

## Discussion

The importance of the hyoid bone and its positioning whether anteriorly or posteriorly to the upper airways has been debated widely in the dental community. However, to the best of our knowledge, no study has been done comparing healthy skeletal Class I and functional activator treated Class II children. Therefore, the aim of this study was to determine the differences, if any, in the position of the hyoid bone in the mentioned groups.

In the perpendicular plane, it was noted that in the present study, the treatment of skeletal Class II malocclusion with an activator resulted in a significant (P<0.01) displacement of the hyoid bone in an anterior and inferior direction.

Findings reported by previous studies, by Battagel *et al*., (1999), Liu *et al*., (2000), Almeida *et al*., (2006), Poon *et al*., (2008) and Doff *et al*., (2009), showed that the hyoid bone is positioned more upwardly after functional appliance therapy. This is a disagreement with the present study, where the hyoid bone takes a more inferior position.

In the present study, the hyoid bone significantly shifts its position to a more anterior one, and the linear distance between hyoidale and the third cervical vertebra is also increased. The linear distance between the mandible from retrognathion and the third cervical vertebra is significantly (P<0.01) increased as well. This anterior displacement present in this study may be due to the functional therapy. Ordubazari *et al*., (1998) disagrees with these results. Other studies agree with the present research (Battagel *et al*.,1999; Zhou *et al*.,2000; Yassei *et al*.,2012)

The present study also revealed that at T2 (roughly a year after the beginning of activator therapy) the hyoid bone continued to be anteriorly displaced. Taking into consideration that the hyoid bone displaces itself in the anterior direction during growth (Ordubazari *et al*.,1998), this anterior shift could be explained with the stability of the lower jaw and an increased ANB changes after functional therapy and stretching of the genioglossus muscle (Yassei *et al*.,2012).

The lowest section of the upper airways as affected by displacement to the anterior direction of the hyoid bone as induced by the activator are also significantly (P<0.01) increased. Of the two newly proposed linear distances, the most inferior one (LspwEb) experiences a significant (P<0.01) increase from T0 to T1 in the Activator group, and the achieved enlargement is kept stable (P<0.05) at T2. This may be explained with the fact that as the activator places the mandible in a more anterior position, the base of the tongue along with the hyoid bone is dragged forward by the muscles.

## Conclusion

The hyoid bone takes a more anterior position when the activator is inserted thus increasing the size of the lowest section of the upper airways. Due to the tendency towards collapsibility of the airways during sleep, a widening of the lower section of the airways could be seen as a positive side effect of continuous use of the Activator appliance. Whether these changes are kept throughout growth and development is a prospect that could be investigated in the future.

## Data Availability

The authors confirm that the data supporting the findings of this study are available within the article [and/or] its supplementary materials.

